# Quantifying absolute treatment effect heterogeneity for time-to-event outcomes across different risk strata: divergence of conclusions with risk difference and restricted mean survival difference

**DOI:** 10.1101/2024.12.19.24319347

**Authors:** Carolien C.H.M. Maas, David M. Kent, Avinash G. Dinmohamed, David van Klaveren

## Abstract

**Background:** Risk-based analyses are increasingly popular for understanding heterogeneous treatment effects (HTE) in clinical trials. For time-to-event analyses, the assumption that high-risk patients benefit most on the clinically important absolute scale when hazard ratios (HRs) are constant across risk strata might not hold. Absolute treatment effects can be measured as either the risk difference (RD) at a given time point or the difference in restricted mean survival time (ΔRMST) which aligns more closely with utilitarian medical decision-making frameworks. We examined risk-based HTE analyses strata in time-to-event analyses to identify the patterns of absolute HTE across risk strata, and whether ΔRMST may lead to more meaningful treatment decisions than RD.

**Methods:** Using artificial and empirical time-to-event data, we compared RD—the difference between Kaplan-Meier estimates at a certain time point—and ΔRMST—the area between the Kaplan-Meier curves—across risk strata and show how these metrics can prioritize different subgroups for treatment. We explored scenarios involving constant HRs while varying both the overall event rates and the discrimination of the risk models.

**Results:** When event rates and discrimination were low, RD and ΔRMST increased monotonically, with high-risk patients benefitting more than low-risk patients. As the event rate increased and/or discrimination increased: 1) a “sweet spot” pattern emerged: intermediate-risk patients benefit more than low-risk and high-risk patients; and 2) RD understates the benefit in high-risk patients.

**Conclusions:** The pattern of HTE characterized by RD may diverge substantially from ΔRMST, potentially leading to treatment mistargeting. Therefore, we recommend ΔRMST for assessing absolute HTE in time-to-event data.

**Key messages:**

1. To quantify absolute heterogeneous treatment effect (HTE) in time-to-event data, the difference in restricted mean survival time (ΔRMST) is more intuitive and comprehensive, less dependent on the time horizon, and better captures HTE when the hazard ratio (HR) of treatment varies over time, compared to the risk difference (RD).
2. We examined risk-based HTE analyses in time-to-event analyses to identify the patterns of absolute HTE across different risk strata, and whether ΔRMST may lead to more meaningful treatment decisions than RD.
3. Even with a constant HR, intermediate-risk patients may benefit more than low-risk and high-risk patients as event rates increase, a phenomenon known as a “sweet spot” pattern.
4. The RD does not accurately reflect the benefit for high-risk patients when event rates and/or discrimination of the risk model are high, unlike to the ΔRMST.
5. We recommend the ΔRMST for assessing absolute HTE, as the RD may potentially lead to treatment mistargeting.

## Introduction

Treatment efficacy is usually assessed by the average outcome difference between two treatments. However, the treatment effect differs among patient groups with varying characteristics.^1–6^ To assess heterogeneous treatment effects (HTE), one-variable-at-a-time subgroup analysis has well-established limitations: it does not account for multiple characteristics simultaneously, it is vulnerable to false positive results due to multiple comparisons, and it has inadequate power to detect true differences.^7^ Predictive approaches to HTE analyses, whereby a model is used to estimate individualized treatment effects taking into account multiple variables, has emerged as an alternative approach to address some of these limitations, including “risk modelling”, which evaluates HTE across strata, defined by a multivariable risk score.^7^

Analyses are often conducted on a relative scale for computational convenience. For instance, the odds ratio (OR) is typically estimated using logistic regression, while the hazard ratio (HR) is calculated using the Cox proportional hazards model. To support clinical decision-making, variation in treatment effects should be evaluated on an absolute scale because they need to be weighed against the adverse effects and costs of treatment.^1^ The ultimate goal of HTE analysis is to evaluate whether the absolute benefits across subgroups vary, such that they exceed the adverse effects and costs for some patients but not others. Whether or not relative effects are approximately similar across risk strata, absolute effects may vary to a clinically meaningful degree—with high-risk groups often (but not always) getting substantially more benefit than lower-risk groups.^8^

When the relative treatment effect is constant across risk strata, it is generally assumed that absolute treatment benefits increase with higher risk. However, if the OR or HR for treatment remains constant across risk strata for binary or time-to-event outcomes, respectively, this does not lead to monotonically increasing absolute benefits (Figure 1; Supplemental Information).^2^ Therefore, absolute effects must be explicitly estimated. The most frequently used approach to evaluating absolute effects is to examine the risk difference (RD)—the difference in estimates of the probability of the outcome of interest between two treatments at a certain time point. For binary outcomes, RD is estimated in each risk quarter by the difference in the outcome rate between the two treatments at a certain time point. When survival times are censored, the difference between the Kaplan-Meier estimates of the two treatments at a certain time point is used to estimate RD.

**Figure 1.**
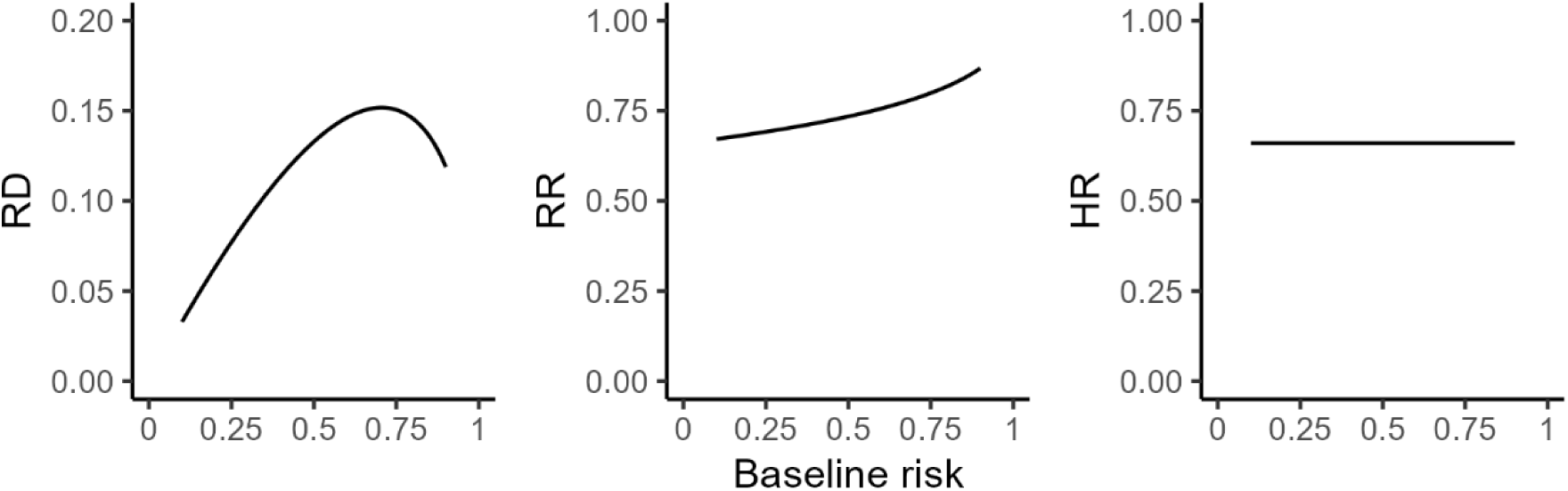
An illustration of the scale dependence of HTE. When treatment effect is non-null and baseline risk varies, HTE is inevitable on at least two out of three of the most commonly used scales for treatment effect. The graphs above show held the hazard ratio (HR=0.66) is held constant, the risk difference (RD) and the relative reduction (RR) are not constant across baseline risk. Results are shown over baseline outcome risks ranging from 0.1 to 0.9. The Supplemental Information offers further details on the definitions of the scales used.

In time-to-event data, the RD heavily depends on the time point chosen, especially, when the hazards are not proportional—i.e., the HR varies over time— and even more so when the Kaplan-Meier curves cross. An alternative approach to quantifying absolute treatment effects in time-to-event data is to estimate the difference in restricted mean survival time (ΔRMST), i.e., the difference in life expectancy between two treatment regimens during a defined period.^9^ RMST can be estimated by the area under the Kaplan-Meier curve up to a certain time point. ΔRMST remains valid and interpretable regardless of whether the proportional hazards assumption holds, making it a recommended measure for assessing treatment effects in time-to-event analyses.^9–12^ Furthermore, ΔRMST aligns more closely with the utilitarian framework of medical decision-making than RD since it accounts for differences in benefits over time, not just at a single, often arbitrary time point.

Naturally, ΔRMST can also be used to measure absolute HTE across risk strata of time-to-event data. However, the settings in which ΔRMST may provide different insights compared to RD remain uncertain. Here, we examined risk-based HTE analyses in time-to-event analyses to better understand: 1) the patterns of absolute HTE across different risk strata; and 2) if ΔRMST may lead to more meaningful treatment decisions than RD. For simplicity, we build intuition by focusing on patterns that emerge with a constant HR of treatment across time, but also consider empirical examples in which the proportional hazards assumption does not hold.

## Methods

First, we simulated event times for one million patients (50% treated) using a Weibull distribution with a shape parameter of two, i.e., increasing failure rate. The hazard was modelled using a baseline hazard of 0.15, HR of standard normally distributed risk of exp(0.5), and a proportional HR for treatment of 0.8. Patients were stratified into four risk quarters according to the data generating risk model. We evaluated RD and ΔRMST in these quarters at four time points: 3, 5, 10, and 15 years. The discriminative ability of the data generating risk model was 0.72. Secondly, we varied the event rate by altering the baseline hazard and risk model performance by adjusting the HR of risk. To span nine realistic scenarios, we used average event rates of 10%, 30% and 50% (representing low, medium and high event rates) and risk models stratifying patients into risk quarters with C-indexes of 0.6, 0.75 and 0.9 (representing low, medium and high discrimination).^8, 13^ We quantified the absolute treatment effect by the 10-year RD and 10-year ΔRMST in each risk quarter. We performed sensitivity analyses by generating the event times of patients using a proportional HR for treatment of 0.5 and by using an exponential distribution (i.e., constant failure rate).

We examined risk-stratified RD and ΔRMST in previously analysed randomized clinical trials (RCTs) with non-constant HR for treatment over time and potentially non-constant HR for treatment across risk strata.^8^ The SOLVD Prevention trial (from 1992 with 4228 participants) and SOLVD Intervention trial (from 1991 with 2569 participants) assessed the effectiveness of enalapril versus placebo on heart failure outcomes and all-cause mortality (Supplemental Table 1).^14^ We calculated RD and ΔRMST across four risk quarters, defined by a risk model including pre-established risk factors.

RD and ΔRMST can be easily estimated with the widely used “survival” R-package. All statistical analyses were performed using R statistical software version 4.4.1, and the code was made available at https://github.com/CHMMaas/TutorialRDvsdRMST.

## Results

### Artificial data

At early time points (3 and 5 years), where event rates were low, RD increased monotonically with increasing risk (Figure 2, panel A-B). For intermediate time points (10 years) and event rates, a “sweet spot” pattern emerged according to RD, i.e., intermediate-risk patients benefit more than low-risk and high-risk patients (Figure 2, panel C).^15^ At a later time point (15 years), when event rates were high across each risk quarter, RD decreased with increasing risk (Figure 2, panel D). Although the same effects were characterized by ΔRMST, this pattern was “phase delayed”. Meaning, ΔRMST was gradually increasing with increasing risk at early time points (3, 5, and 10 years), but only at 15 years ΔRMST did it show a “sweet spot” pattern (Figure 2).

**Figure 2.**
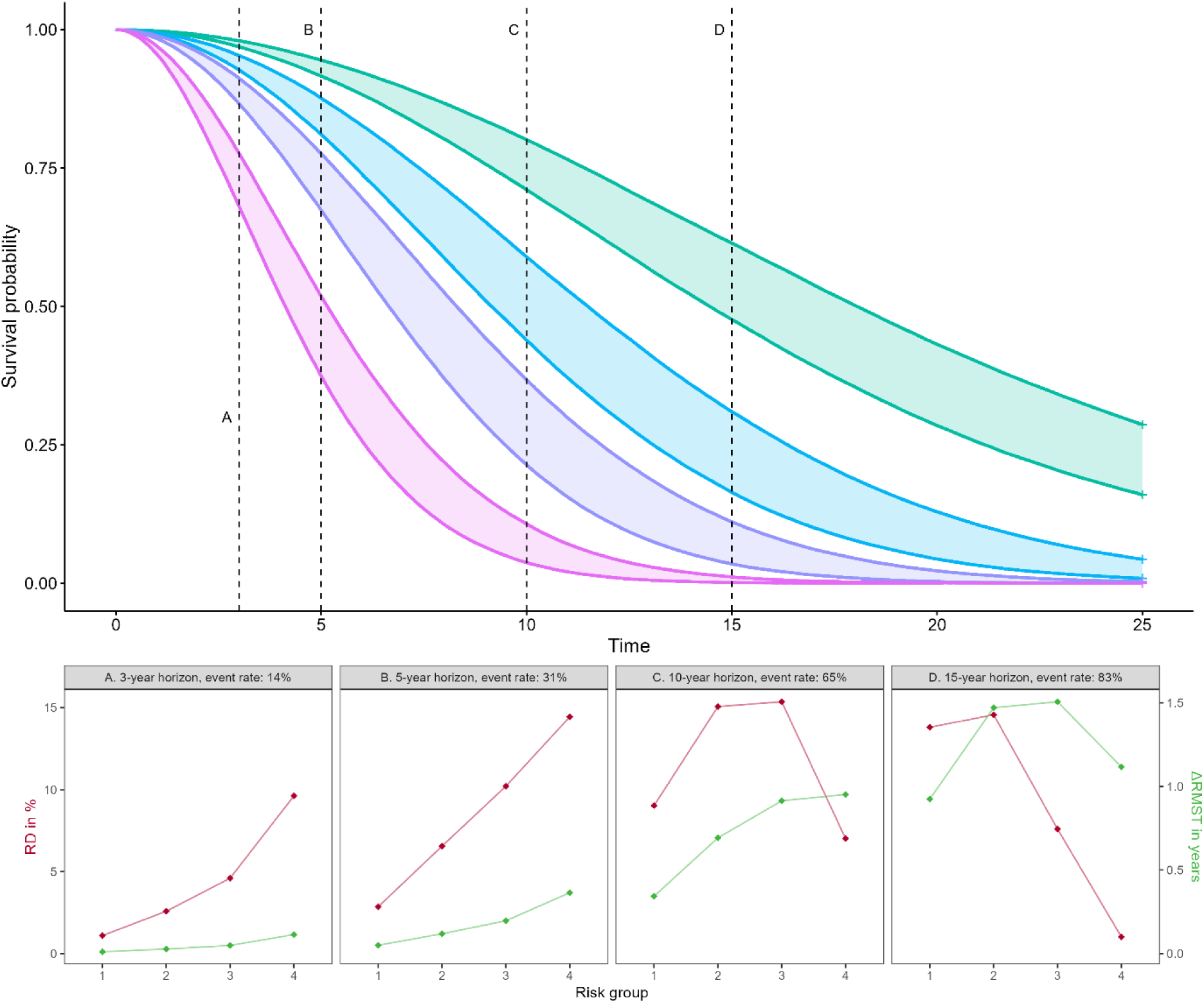
Illustration using artificial data: the pattern of absolute heterogeneous treatment effect (HTE) for the risk difference (RD) may diverge substantially from the difference in restricted mean survival time (ΔRMST) In the top part of the Figure, treatment-stratified survival curves with increasing failure rates are presented for one million patients using a constant overall proportional HR for treatment of 0.8 across the four risk quarters generated using a risk model with a C-index of 0.72. In each pair of colored survival curves, the lower survival curve denotes the treated group, while the upper survival curve represents the control group. Below, panels A-D illustrate the risk difference (ARD, i.e., the difference in survival probabilities) and the difference in restricted mean survival time (ΔRMST, i.e., the area between the survival curves) at four distinct time points.

When the average event rate was low (event rate=10%), both RD and ΔRMST exhibited a comparable pattern of absolute HTE across risk strata, with a gradual increased in benefit as the risk level rose, regardless of the risk models’ discriminative ability (Figure 3A-C). When the discriminative ability of the risk model was low (C-index=0.60), RD and ΔRMST gradually increased with increasing risk, irrespective of the event rate (Figure 3A, D, G). Finally, when the event rate and medium discrimination (event rate=30%; C-index=0.75), the absolute HTE increased monotonically, regardless of the metric (Figure 3E).

**Figure 3.**
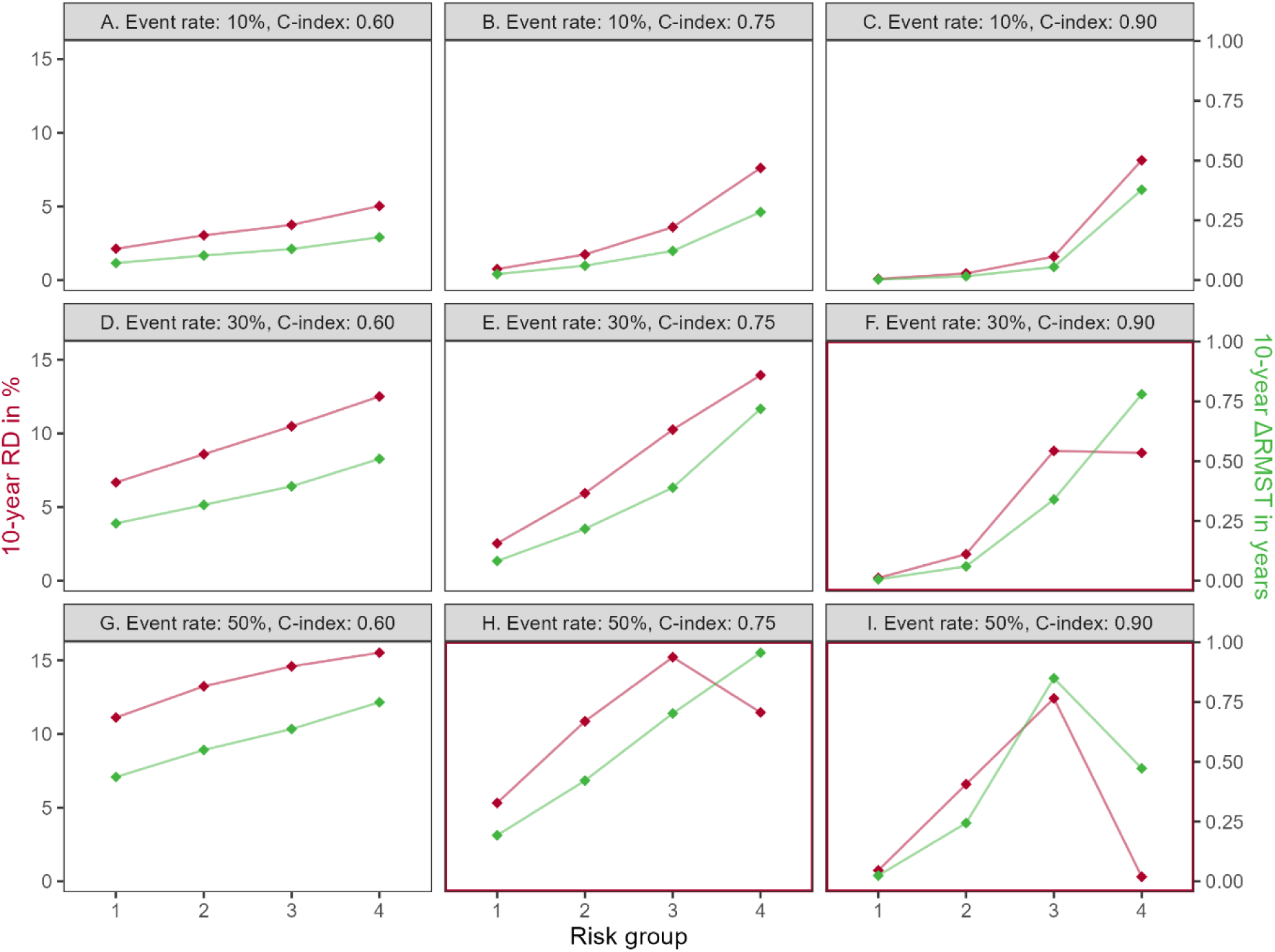
Illustration using artificial data: the difference in restricted mean survival time (ΔRMST) and risk difference (RD) give a similar pattern of absolute treatment effect across risk strata. The event times were generated using increasing failure rates of one million patients in four risk strata, using a constant overall proportional HR for treatment of 0.8, an average event rate of 10%, 30%, and 50% among the control group, and a discriminative ability of the risk-stratified model of 0.60, 0.75, and 0.90. The red highlighted panels indicate where ARD and ΔRMST provide discordant treatment targeting.

Differences in the pattern of absolute HTE between RD and ΔRMST emerged when either event rates or the risk models’ discriminative ability increase. When generating data using a high event rate and medium discrimination (event rate=50%; C-index=0.75) or medium event rate and high discrimination (event rate=30%; C-index=0.90), RD began to display a “sweet spot” pattern, with the third risk quarter having greater benefit than the highest risk quarter—diverging from ΔRMST which still showed a monotonic increase (Figure 3F, H). When generating data using a high even rate and high discrimination (event rate=50%; C-index=0.90), RD and ΔRMST both showed a “sweet spot” pattern, although RD in the highest risk strata reduced to zero whereas ΔRMST remained substantial (Figure 3I).

When the overall treatment effect was larger (HR for treatment was 0.5), the divergence between RD and ΔRMST was observed at higher event rates and higher discrimination of the risk model (Supplemental Figure 1). Similar divergence patterns between RD and ΔRMST were observed using an exponential distribution to generate patient event times (Supplemental Figure 2-3).

### Case studies

At the 90^th^ percentile of follow-up, survival probabilities were 69.5% at 4.6 years (SOLVD Prevention) and 54.5% at 4.3 years (SOLVD Intervention), with discriminative abilities of 0.65 and 0.67, respectively. The RD and ΔRMST demonstrated different patterns across risk strata (Figure 4-5). In the SOLVD Prevention trial, the Kaplan-Meier curves intersected (i.e., non-proportional HR of treatment) within the fourth risk quarter. As a result, the 5-year RD in the fourth risk quarter was negative, while the 5-year ΔRMST remained substantially positive (Figure 4). Similar behaviour in the SOLVD Intervention trial was presented in the second risk quarter (Figure 5).

**Figure 4.**
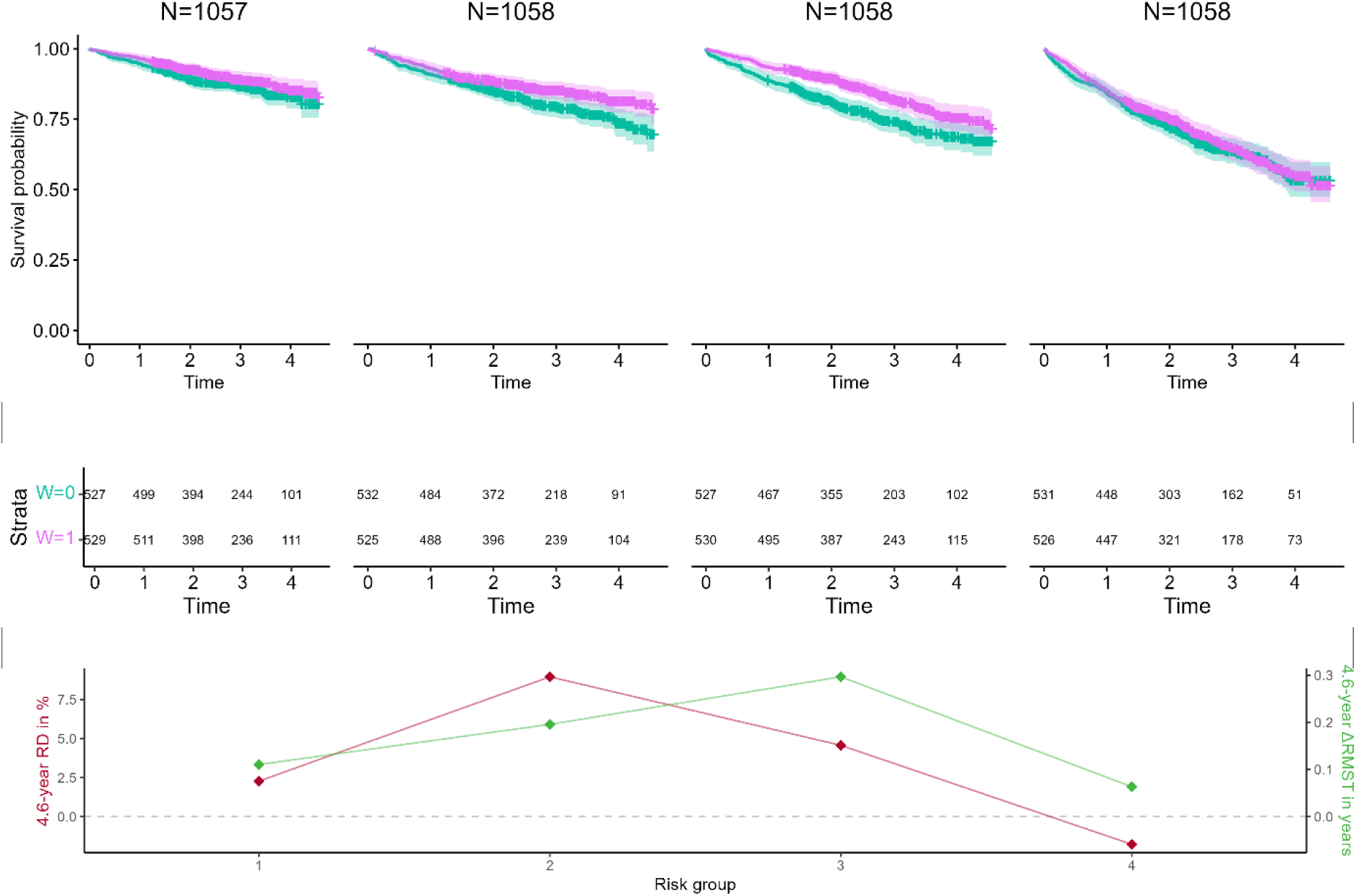
Illustration using empirical data: the difference in restricted mean survival time (ΔRMST) is a more informative measure of absolute treatment effect than the risk difference (RD) due to non-proportional hazards in the SOLVD Prevention trial for death or hospitalization for heart failure. The survival curves were stratified by placebo treatment (pink line) and enalapril treatment (green line) and displayed with 95% confidence intervals. We restricted follow-up to 4.6 years, which is the 90th percentile of follow-up time. The hazard ratio for treatment varied across risk groups: 0.78 (95% CI: 0.44-1.12) in group 1, 0.71 (95% CI: 0.42-0.99) in group 2, 0.67 (95% CI: 0.41-0.93) in group 3, and 0.95 (95% CI: 0.75-1.15) in group 4.

**Figure 5.**
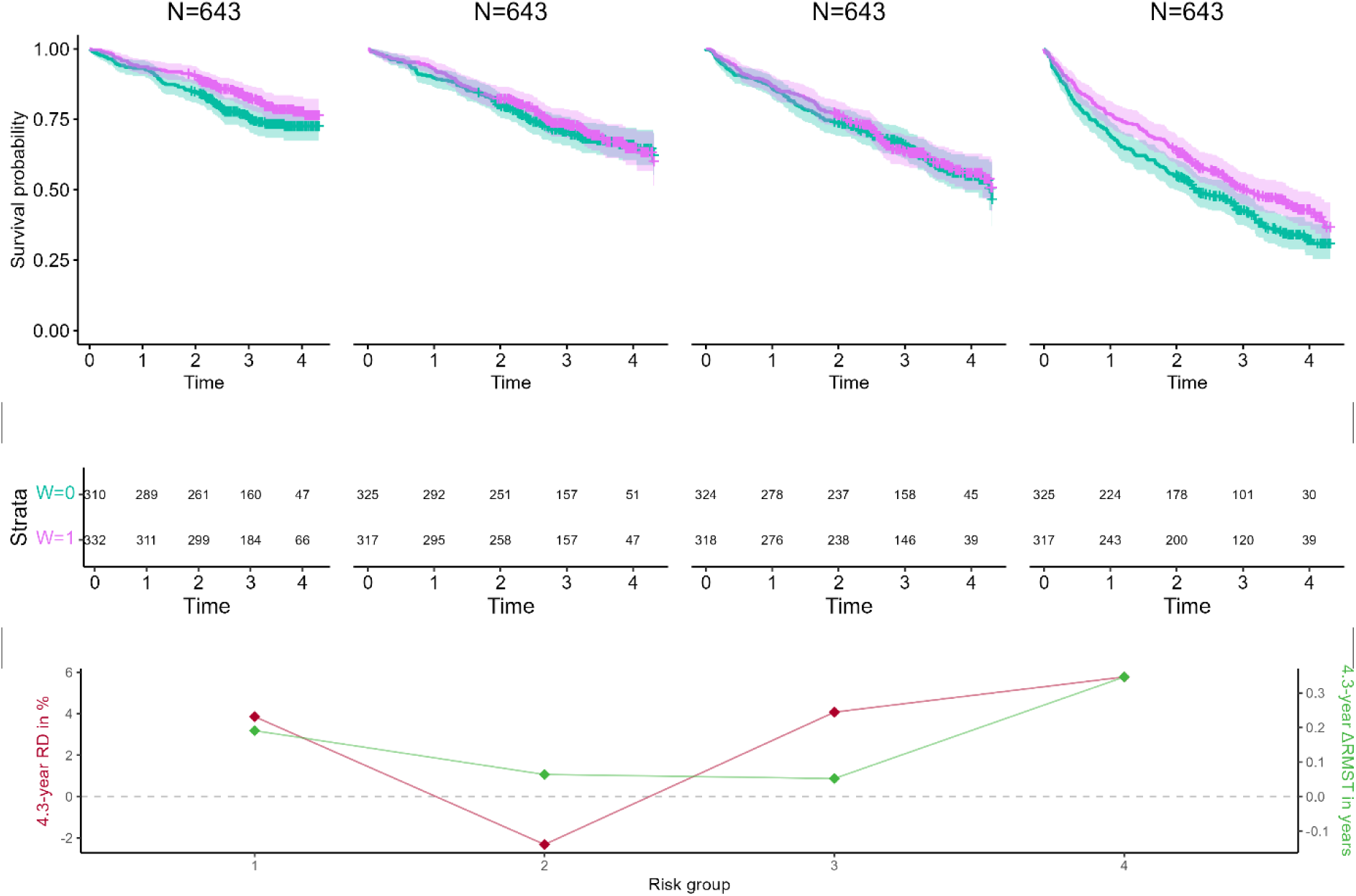
Illustration using empirical data: the difference in restricted mean survival time (ΔRMST) gives similar absolute treatment effect than the risk difference (RD) across risk strata for the outcome hospitalization for worsening heart failure in the SOLVD Intervention trial. The survival curves were stratified by placebo treatment (pink line) and enalapril treatment (green line) and displayed with 95% confidence intervals. We restricted follow-up to 4.6 years, which is the 90th percentile of follow-up time. The hazard ratio for treatment varied across risk groups: 0.74 (95% CI: 0.41-1.07) in group 1, 0.95 (95% CI: 0.66-1.23) in group 2, 0.96 (95% CI: 0.72-1.21) in group 3, and 0.77 (95% CI: 0.56-0.97) in group 4.

## Discussion

It is often assumed that under a constant relative treatment effect, the absolute benefits of treatment increase with increasing risk. This relationship is often assumed in the evidence-based medicine literature and it is implicit in many clinical guidelines that reserve treatments with high costs or adverse effect for those with higher baseline risk.^16, 17^ However, when modelling time-to-event data, it is not broadly appreciated that a constant HR does not yield monotonically increasing predictions of absolute benefit across risk groups. The pattern is influenced by the distribution of predicted risk. A “sweet spot” pattern emerges when a considerable portion of the risk distribution falls below 50%, while another considerable portion exceeds 50%.^15^ Moreover, our findings illustrated that the pattern of absolute HTE can differ substantially when quantified using either ΔRMST or RD. Since ΔRMST aligns more closely with the utilitarian framework of medical decision-making than RD—as it accounts for differences in benefits across time, not at just a single, often arbitrary time point—our findings illustrated that RD may not provide the best representation of differential treatment effects, potentially leading to treatment mistargeting. As seen in our analysis of the SOLVD studies, when the HR of treatment was not constant across risk strata and time, ΔRMST better captures absolute HTE than RD. Thus, we recommend ΔRMST for assessing absolute HTE in time-to-event data.

### Clinical Decision Making

Clinically important HTE is found when absolute HTE estimates span important decision thresholds. Using RD versus ΔRMST might influence decision-making in either a shared decision-making context or a rationing context. For example, presenting the results shown in Figure 3I framed as RD, the benefits of treatment in risk quarters one and four might not be worthwhile, even if the adverse effects and costs of treatment were fairly small—whereas benefits presented by ΔRMST may seem quite worthwhile for risk quarter four. In the context of scarcity and rationing, for example, where the health system can treat only the highest benefit quarter of patients, using the RD scale would target risk quarter three in Figure 3F and 3H, whereas ΔRMST scale would target risk quarter four. Decision making would differ more if HTE was estimated on the individual patient level, rather than at the risk quarter level.

While it may seem evident that RD understates the absolute benefits over time when event rates are high, we have observed that ΔRMST is used only infrequently to characterize absolute benefits in risk modelling studies. In a systematic review of predictive HTE analyses in clinical trials, which referenced the PATH statement for guidance, only one out of 16 studies that examined HTE using time-to-event data with a risk modelling approach utilized ΔRMST as a measure of absolute HTE.^16, 18^ In the remaining 15 studies, ΔRMST was not used to assess absolute HTE, despite high event rates. In one of these analyses, the investigators hypothesized that the lack of relative HTE would lead to monotonically increasing absolute benefits even though the overall event rate was near 50%.^19^

### Limitations to this study

This study has several limitations. First, the artificial trial data does not fully capture scenarios that could be encountered in the real world. To build intuition, we made two assumptions: 1) the HR for treatment is constant across risk strata, and 2) the HR for treatment remains constant over time. Concerning the first assumption, if the HR for treatment would vary across risk strata, we still expect ΔRMST to provide a better reflection of absolute HTE compared to RD. Regarding the second assumption, we focused on the simple case of proportional hazards, even though it has been pointed out that the proportional hazards assumption is causally implausible since susceptible individuals are differentially depleted from the treatment arms over time.^20, 21^ Nevertheless, we think this is useful for illustrative purposes, noting that an empirical investigation of 27 trials failed to detect evidence for this selection bias across time, suggesting proportional hazards might often provide a reasonable approximation of effects.^22^ We also focused on “risk modelling” but note that “effect modelling” is increasingly popular.^16^ ΔRMST can be applied to HTE analyses where patients are stratified based on benefit rather than risk^23^, although the best methods for reliable effect modelling are still emerging.

### Advantages and disadvantages of using ΔRMST to measure absolute HTE

Prior studies have emphasized that the advantage of using ΔRMST over HR becomes evident when the proportional hazards assumption is substantially violated.^9, 11, 12, 24–26^ In this analysis, we highlighted the benefits of ΔRMST over RD. First of all, ΔRMST may be more intuitive for patients and clinicians as it measures the difference in the average time until an event occurs between two treatment groups within a specified period.^26^ Most importantly, ΔRMST provides a more comprehensive measure of treatment effect by considering the entire survival curve up to a specified time point. Choosing an appropriate time horizon is needed for both ΔRMST and RD. Tian et al. demonstrated that appropriate statistical inference is possible when estimating RMST up to the longest follow-up time when this is clinically useful.^27^ In contrast, RD estimates at the longest follow-up can be unstable due to fewer individuals remaining. Thus, ΔRMST is particularly useful when there is a high loss to follow-up, such as near the maximum follow-up time. Lastly, when the event rate exceeds 50%, ΔRMST is more advantageous than RD, as RD understates the benefit. The divergence between RD and ΔRMST might be less evident in RCT data due to strict inclusion criteria, leading to healthier, more homogeneous participants, which results in lower event rates and reduced risk model discrimination. Observational studies often have higher event rates, such as in frail patients or those with aggressive cancers. While our analyses focused on RCT data, ΔRMST can also quantify absolute HTE in observational data when properly addressing treatment group imbalances, as in a target trial emulation framework.^28–30^

Nevertheless, ΔRMST shares some of the same limitations as RD. Its estimates are confined to the trial’s duration, making long-term effects hard to infer from short trials.^26^ Additionally, its sensitivity to follow-up time complicates comparisons of intervention efficacy or safety across studies with varying follow-up duration and event rates.

### Conclusion

Our findings highlight the limitations of RD in risk-based HTE analyses for time-to-event outcomes. We recommend using ΔRMST to assess absolute HTE for time-to-event outcomes.

## Data Availability

The trial data can be requested at NHLBI (https://biolincc.nhlbi.nih.gov/studies). Project home page: https://github.com/CHMMaas/TutorialRDvsdRMST.

## Abbreviations

RD: Risk difference
ATE: Average treatment effect
ΔRMST: Difference in restricted mean survival time
HR: Hazard ratio
HTE: Heterogeneous treatment effect

## Declarations

### Ethics approval

No ethics approval was needed for this research.

### Data availability

The trial data can be requested at NHLBI (https://biolincc.nhlbi.nih.gov/studies).

Project home page: https://github.com/CHMMaas/TutorialRDvsdRMST.

### Author contributions

C.C.H.M. Maas: Conceptualization, Methodology, Software, Formal analysis, Writing – Original Draft. D.M. Kent: Conceptualization, Writing – Review & Editing. A.G. Dinmohamed: Conceptualization, Writing – Review & Editing. D. van Klaveren: Conceptualization, Methodology, Writing – Review & Editing.

## References

1. Kravitz RL, Duan N Fau - Braslow J, Braslow J. Evidence-based medicine, heterogeneity of treatment effects, and the trouble with averages. 2004.

2. Dahabreh IJ, Hayward R, Kent DM. Using group data to treat individuals: understanding heterogeneous treatment effects in the age of precision medicine and patient-centred evidence. Int J Epidemiol 2016; 45: 2184–93.

3. Davidoff F. Can Knowledge About Heterogeneity in Treatment Effects Help Us Choose Wisely? 2017.

4. Hayward RA, Kent Dm Fau - Vijan S, Vijan S Fau - Hofer TP, Hofer TP. Reporting clinical trial results to inform providers, payers, and consumers. 2005.

5. Kent DM, Rothwell Pm Fau - Ioannidis JPA, Ioannidis Jp Fau - Altman DG, Altman Dg Fau - Hayward RA, Hayward RA. Assessing and reporting heterogeneity in treatment effects in clinical trials: a proposal. 2010.

6. Sun X, Ioannidis JP, Agoritsas T, Alba AC, Guyatt G. How to use a subgroup analysis: users’ guide to the medical literature. 2014.

7. Kent DM, Steyerberg EW, van Klaveren D. Personalized evidence based medicine: predictive approaches to heterogeneous treatment effects. The BMJ 2018; 363.

8. Kent DM, Nelson J, Dahabreh IJ, Rothwell PM, Altman DG, Hayward RA. Risk and treatment effect heterogeneity: re-analysis of individual participant data from 32 large clinical trials. International Journal of Epidemiology 2016.

9. Royston P, Parmar MK. The use of restricted mean survival time to estimate the treatment effect in randomized clinical trials when the proportional hazards assumption is in doubt. Stat Med 2011; 30: 2409–21.

10. Kim DH, Shi SM, Carroll D, Najafzadeh M, Wei LJ. Restricted mean survival time versus conventional measures for treatment decision-making. J Am Geriatr Soc 2021; 69: 2282–9.

11. A’Hern RP. Restricted Mean Survival Time: An Obligatory End Point for Time-to-Event Analysis in Cancer Trials? J Clin Oncol 2016; 34: 3474–6.

12. Trinquart L, Jacot J, Conner SC, Porcher R. Comparison of Treatment Effects Measured by the Hazard Ratio and by the Ratio of Restricted Mean Survival Times in Oncology Randomized Controlled Trials. J Clin Oncol 2016; 34: 1813–9.

13. Wessler BS, Nelson J, Park JG, et al. External Validations of Cardiovascular Clinical Prediction Models: A Large-Scale Review of the Literature. Circ Cardiovasc Qual Outcomes 2021; 14: e007858.

14. Investigators S, Yusuf S, Pitt B, Davis CE, Hood WB, Jr., Cohn JN. Effect of enalapril on mortality and the development of heart failure in asymptomatic patients with reduced left ventricular ejection fractions. N Engl J Med 1992; 327: 685–91.

15. Redelmeier DA, Thiruchelvam D, Tibshirani RJ. Testing for a Sweet Spot in Randomized Trials. Medical Decision Making 2022; 42: 208–16.

16. Selby JV, Maas C, Fireman BH, Kent DM. Impact of the PATH Statement on Analysis and Reporting of Heterogeneity of Treatment Effect in Clinical Trials: A Scoping Review. medRxiv 2024.

17. Harrell F. Viewpoints on Heterogeneity of Treatment Effect and Precision Medicine. 2018 2024 [cited 2024; Available from: https://www.fharrell.com/post/hteview/

18. Charu V, Liang JW, Chertow GM, et al. Heterogeneous Treatment Effects of Intensive Glycemic Control on Kidney Microvascular Outcomes and Mortality in ACCORD. Journal of the American Society of Nephrology 2024; 35: 216–28.

19. Bond MJG, van Smeden M, Degeling K, et al. Predicting Benefit From FOLFOXIRI Plus Bevacizumab in Patients With Metastatic Colorectal Cancer. JCO Clin Cancer Inform 2024; 8: e2400037.

20. Hernán MA. The hazards of hazard ratios. Epidemiology 2010; 21: 13–5.

21. Stensrud MJ, Hernán MA. Why Test for Proportional Hazards? Jama 2020; 323: 1401–2.

22. Strobel A, Wienke A, Kuss O. How hazardous are hazard ratios? An empirical investigation of individual patient data from 27 large randomized clinical trials. European Journal of Epidemiology 2023; 38: 859–67.

23. Desai RJ, Glynn RJ, Solomon SD, Claggett B, Wang SV, Vaduganathan M. Individualized Treatment Effect Prediction with Machine Learning - Salient Considerations. NEJM Evid 2024; 3: EVIDoa2300041.

24. Uno H, Claggett B, Tian L, et al. Moving beyond the hazard ratio in quantifying the between-group difference in survival analysis. J Clin Oncol 2014; 32: 2380–5.

25. Kim DH, Uno H, Wei LJ. Restricted Mean Survival Time as a Measure to Interpret Clinical Trial Results. JAMA Cardiol 2017; 2: 1179–80.

26. Kloecker DE, Davies MJ, Khunti K, Zaccardi F. Uses and Limitations of the Restricted Mean Survival Time: Illustrative Examples From Cardiovascular Outcomes and Mortality Trials in Type 2 Diabetes. Ann Intern Med 2020; 172: 541–52.

27. Tian LA-O, Jin HA-O, Uno HA-O, et al. On the empirical choice of the time window for restricted mean survival time. 2020.

28. Rekkas A, van Klaveren D, Ryan PB, Steyerberg EW, Kent DM, Rijnbeek PR. A standardized framework for risk-based assessment of treatment effect heterogeneity in observational healthcare databases. arXiv:201006430v2 2022.

29. Maas CCHM, van Klaveren D, Durmaz M, et al. Comparative effectiveness of 6x R-CHOP21 versus 6x R-CHOP21 + 2 R for patients with advanced-stage diffuse large B-cell lymphoma. Blood Cancer Journal 2024; 14: 157.

30. Hernán MA, Robins JM. Using Big Data to Emulate a Target Trial When a Randomized Trial Is Not Available. Am J Epidemiol 2016; 183: 758–64.

