## Supplemental Material for "Quantifying absolute treatment effect heterogeneity for time-to-event outcomes across different risk strata: divergence of conclusions with risk difference and restricted mean survival difference"

**Supplemental Information. Relationship between the risk difference (RD), relative risk (RR), and hazard ratio (HR) in time-to-event data.**

**Definitions**

**Mortality probability** $p(t) = P(T \leq t)$ represents the probability that a subject will die before time $t$.

**Risk difference** $RD = p(t|control) - p(t|treated)$ is the difference in survival probabilities between the treated and control groups at time $t$.

**Relative risk** $RR = \frac{p(t|treated)}{p(t|control)}$ is the ratio of the mortality probabilities between the treated and control groups at time $t$.

**Hazard ratio** $\mathrm{HR} = \frac{h(t|treated)}{h(t|control)}$ is the ratio of hazards between the treated and control groups at time $t$.

**Constant HR**

When the HR is constant, $h(t) = H(t)$.

Thus, $\mathrm{HR} = \frac{h(t|treated)}{h(t|control)} = \frac{H(t|treated)}{H(t|control)} = \frac{-\ln(S(t|treated))}{-\ln(S(t|control))} = \frac{\ln\left(s(t|treated)\right)}{\ln\left(s(t|control)\right)} = \frac{\ln\left(1-p(t|treated)\right)}{\ln\left(1-p(t|control)\right)}$

Thus, $p(t|treated) = 1 - \left(1 - p(t|control)\right)^{HR}$

Therefore, $RD = p(t|control) - 1 + \left(1 - p(t|control)\right)^{HR}$

And $RR = \frac{1-\left(1-p(t|control)\right)^{HR}}{p(t|control)}$

**Supplemental Table 1. Detailed description of the randomized clinical trials and performance metrics of their internally developed risk model.**

| | SOLVD Prevention | SOLVD Intervention |
|---|---|---|
| Year[1] | 1992[1] | 1991[1] |
| Patient population | Congestive heart failure | Congestive heart failure |
| Treatment | Enalapril versus placebo | Enalapril versus placebo |
| N[2] | Prevention: 4,228 | Intervention: 2,569 |
| Outcome | Death or hospitalization due to heart failure | All-cause mortality |
| Follow-up[3] | 3.1 [2.2; 4.0] | 3.5 [2.9; 4.0] |
| 90th percentile[4] | 4.6 | 4.3 |
| KM[5] | 69.5 [67.4; 71.6] | 54.5 [51.7; 57.4] |
| RD | 0.03 [-0.01; 0.07] | 0.03 [-0.02; 0.09] |
| p-value[6] | 0.001 | 0.009 |
| ΔRMST | 0.16 [0.08; 0.25] | 0.18 [0.06; 0.29] |
| Predictors | Age, blood urea nitrogen, NYHA heart failure class, diuretic therapy, sex, race, history of CABG or other cardiac surgery, history of chronic obstructive pulmonary disease, history of atrial fibrillation, SBP, DBP, heart rate, ACE inhibitor therapy, beta blocker therapy, creatinine, potassium, white blood cell count[2-4] | Age, sex, body weight, heart rate, current smoking status, history of diabetes, history of MI, history of oedema, history of chronic obstructive pulmonary disease, NYHA heart failure class, left ventricular ejection fraction, history of atrial fibrillation ACE inhibitor therapy, beta blocker therapy, creatinine, SBP, DBP, sodium, blood urea nitrogen[5-8] |
| C-index | 0.65 | 0.67 |

[1]Year of publication with references; [2]Sample size for patients with an outcome recorded; [3]Median [IQR] follow-up time calculated using reverse Kaplan-Meier estimates; [4]90th percentile of follow-up time using reverse Kaplan-Meier estimates. [5]KM is the Kaplan-Meier estimate at the 90th percentile of follow-up time with a 95% confidence interval; [6]p-value of the log-rank test to test if the treatment-stratified Kaplan-Meier curves truncated at 90th percentile of follow-up time were significantly different. Abbreviations: ACE = angiotensin-converting-enzyme; CABG = coronary artery bypass graft; DBP = diastolic blood pressure; NYHA = New York Heart Association; MI = myocardial infarction; SBP = systolic blood pressure, ΔRMST = difference in restricted mean survival time.

**Supplemental Figure 1. Illustration using artificial data: the difference in restricted mean survival time (ΔRMST) and risk difference (RD) give a similar pattern of absolute treatment effect across risk strata.** Simulated the event times of 1,000,000 individuals in four risk strata using an increasing failure rate, a constant overall proportional HR for treatment of 0.5, an average overall event rate of 10%, 30%, and 50% among the control group, and a discriminative ability of the risk-stratified model of 0.60, 0.75, and 0.90. The red highlighted panels indicate where RD and ΔRMST provide discordant treatment targeting.

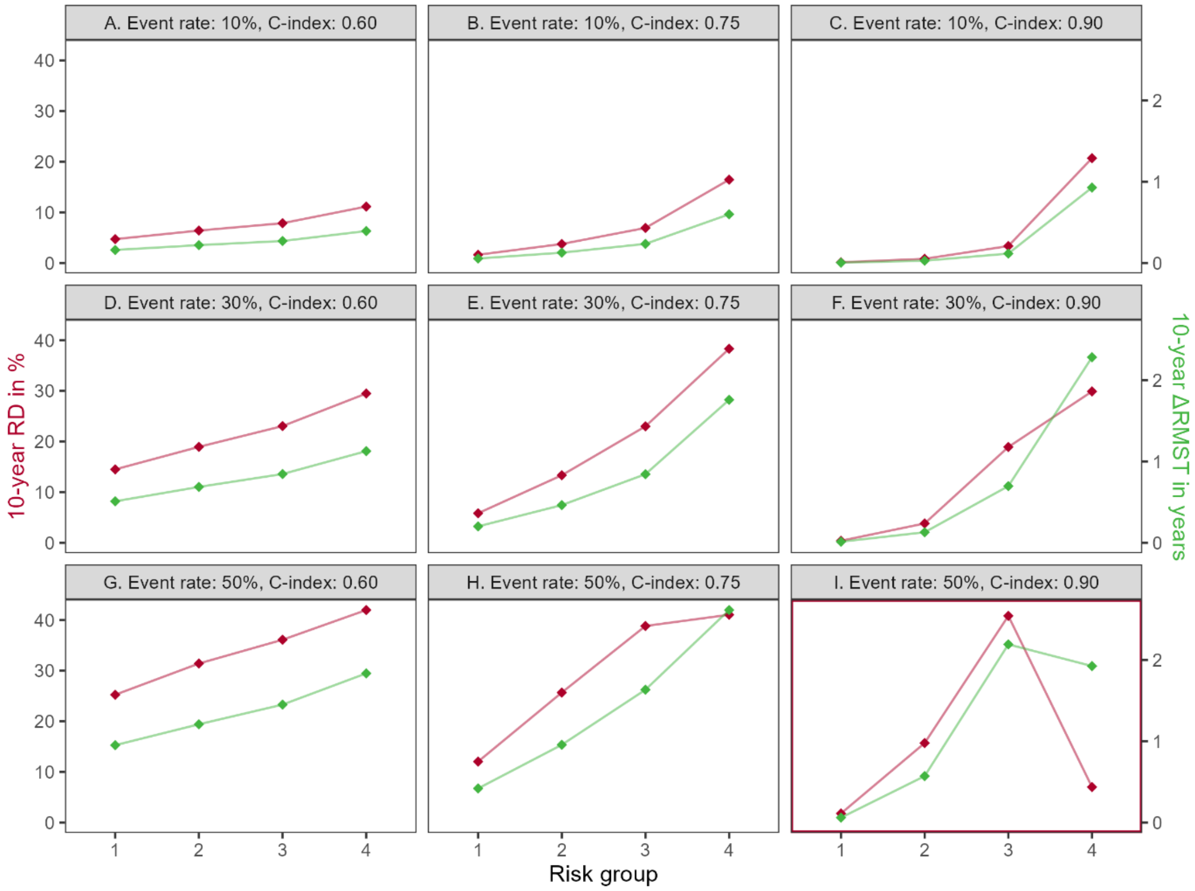

**Supplemental Figure 2. Illustration using artificial data: the difference in restricted mean survival time (ΔRMST) and risk difference (RD) give a similar pattern of absolute treatment effect across risk strata.** Simulated the event times of 1,000,000 individuals in four risk strata using a constant failure rate, a constant overall proportional HR for treatment of 0.8, an average overall event rate of 10%, 30%, and 50% among the control group, and a discriminative ability of the risk-stratified model of 0.60, 0.75, and 0.90. The red highlighted panels indicate where RD and ΔRMST provide discordant treatment targeting.

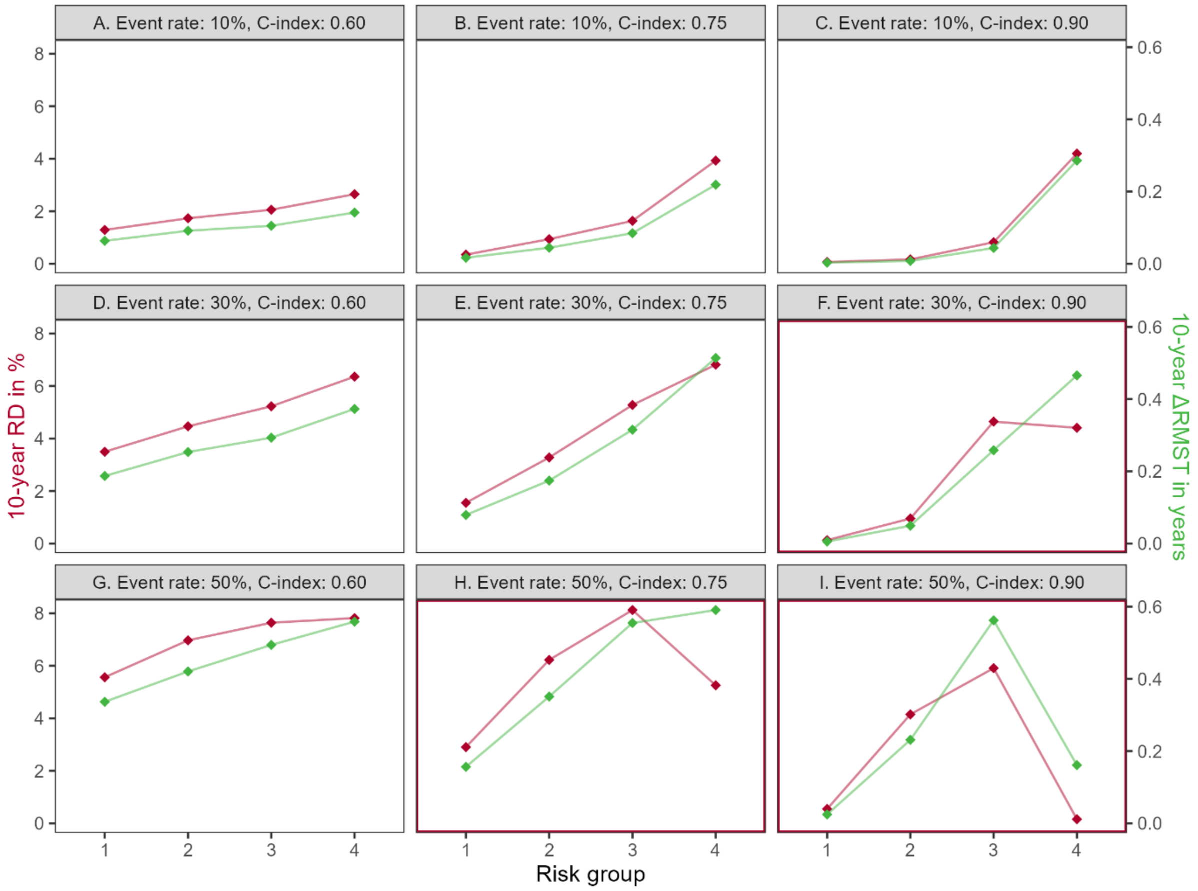

**Supplemental Figure 3. Illustration using artificial data: the difference in restricted mean survival time (ΔRMST) and risk difference (RD) give a similar pattern of absolute treatment effect across risk strata.** Simulated the event times of 1,000,000 individuals in four risk strata using a constant failure rate, a constant overall proportional HR for treatment of 0.5, an average overall event rate of 10%, 30%, and 50% among the control group, and a discriminative ability of the risk-stratified model of 0.60, 0.75, and 0.90. The red highlighted panels indicate where RD and ΔRMST provide discordant treatment targeting.

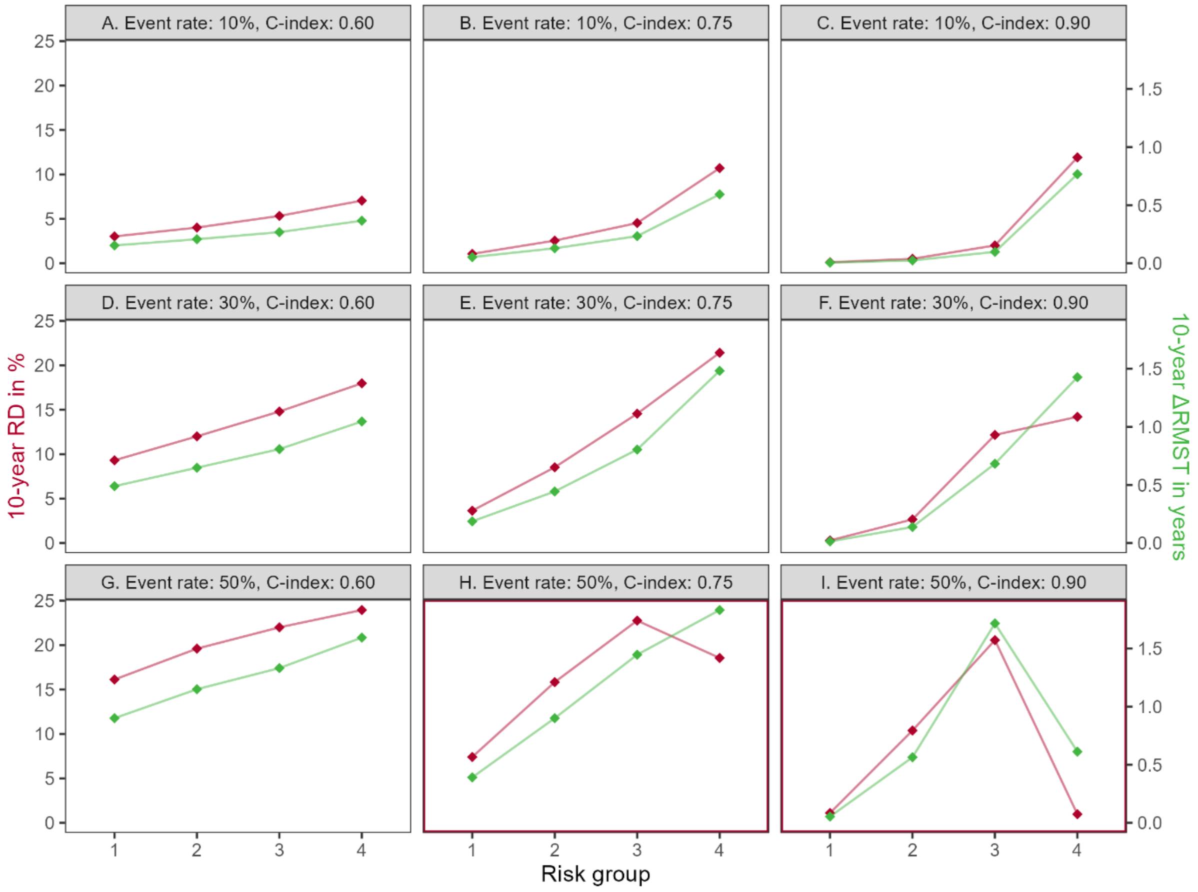

**References**

1. Investigators S, Yusuf S, Pitt B, Davis CE, Hood WB, Jr., Cohn JN. Effect of enalapril on mortality and the development of heart failure in asymptomatic patients with reduced left ventricular ejection fractions. *N Engl J Med* 1992; **327**: 685-91.
2. Abraham WT, Fonarow GC, Albert NM, et al. Predictors of in-hospital mortality in patients hospitalized for heart failure: insights from the Organized Program to Initiate Lifesaving Treatment in Hospitalized Patients with Heart Failure (OPTIMIZE-HF). *J Am Coll CRDiol* 2008; **52**: 347-56.
3. O'Connor CM, Abraham WT, Albert NM, et al. Predictors of mortality after discharge in patients hospitalized with heart failure: an analysis from the Organized Program to Initiate Lifesaving Treatment in Hospitalized Patients with Heart Failure (OPTIMIZE-HF). *Am Heart J* 2008; **156**: 662-73.
4. Levy WC, Mozaffarian D, Linker DT, et al. The Seattle Heart Failure Model: prediction of survival in heart failure. *Circulation* 2006; **113**: 1424-33.
5. Bouvy ML, Heerdink ER, Leufkens HG, Hoes AW. Predicting mortality in patients with heart failure: a pragmatic approach. *Heart* 2003; **89**: 605-9.
6. Lee DS, Austin PC, Rouleau JL, Liu PP, Naimark D, Tu JV. Predicting mortality among patients hospitalized for heart failure: derivation and validation of a clinical model. *Jama* 2003; **290**: 2581-7.
7. Pocock SJ, Wang D, Pfeffer MA, et al. Predictors of mortality and morbidity in patients with chronic heart failure. *Eur Heart J* 2006; **27**: 65-75.
8. Senni M, Santilli G, Parrella P, et al. A novel prognostic index to determine the impact of cRDiac conditions and co-morbidities on one-year outcome in patients with heart failure. *Am J CRDiol* 2006; **98**: 1076-82.
